# Effect of mangrove-sword bean-food bar on weight and Weight-for-Age Z-score in under-five children after landslide disaster

**DOI:** 10.1101/2023.06.01.23290845

**Authors:** Fatmah, Suyud Warno Utomo

## Abstract

Special attention needs to be given to under-five children who have specific needs due to their lower ability to prepare for disasters. Simple, ready-to-eat, and tasty food that meets the micronutrient needs of the groups is needed. The study aimed to assess the effect of the food bar made from *api-api* mangrove (*Avicennia marina*) and sword bean (*Canavalia ensiformis*) blends on the weight and Weight-for-Age Z (WAZ) score after landslide disaster. A non randomized pre-post intervention study was performed on 34 under-five children affected by landslide disaster during 15 days. Subjects were divided into intervention (a*pi-api* mangrove-*sword bean* food bars) and control groups (sword bean food bars). Both groups received the food bars for 15 days. Education on balanced nutrition for under-five children was provided to all subjects’ mothers in the study. The study revealed that most mothers of under-five children liked mangrove-sword bean food bar from the aroma, taste, texture, and color in the hedonic test. The intervention group experienced weight gain (0.32 kg) and WAZ score (0.73) were greater compared with the control group. There were significant difference in weight and WAZ score changes of the under-five children accompanied by a significant increases in mothers’ knowledge on the balanced nutrition for under-five children and food bar total consumption. No significant differences of macronutrient consumption, except fat intake between pre-post study were found. *Api-api* mangrove-*sword bean* food bar can be an emergency food alternative for disaster vulnerable group communities affected by natural disasters. Future studies may wish to consider examining the issue using pregnant women as the research subjects.

## Introduction

A continuous increase in hydrometeorological disasters, such as earthquakes, tsunamis, floods, and landslides, is currently observed in Indonesia, which is partly caused because the country is located between two large oceans and on the meeting points of the world’s three major plates (Indo-Australia, Eurasia, and Pacific plates). The diverse land surface reliefs of this country also contribute to this vulnerability to disasters. As of December 2022, 3,318 disasters have struck Indonesia, with floods as the most frequent disasters (42.8%) [1].

West Java Province, the most populated province in Indonesia, ranks first among the provinces with the highest incidences of natural disasters in Indonesia from January to November 2022 [2]. Sumedang District, one of the districts in this province, experienced 85 landslides in 2021 alone [3]. In January 2022, a landslide occurred after the area experienced high-intensity rain that lasted 2 hours. This landslide claimed 40 lives while severely damaging many buildings [4].

One of the main problems faced in natural disaster management is fulfilling nutritional needs, especially for vulnerable groups, including under-five children. On the other hand, under-five children often experience malnutrition during a disaster caused by a low intake of macronutrients. Under-five children need age-appropriate balanced nutrients. To overcome this issue, Emergency Food Products (EFP) with complete nutrients should be able to be consumed directly as the main source of energy for 15 days during the disaster management period or start from the evacuation time [5-6].

Several emergency food formulations have been developed in Indonesia, including food bars made from brand and corn flours [7], cookies from Moringa leaf flour [8], cookies from sweet potato flour, banana flour, mung bean flour [9], and soybean broccoli mangrove fruit food bar [10]. However, even though *api-api* mangrove (Avicennia marina) is considered one of the ingredients that can substitute rice and wheat flour for making biscuits, it has never been used to make food bars. The *api-api* mangrove has a higher energy content than rice and corn [11]. However, the fat, carbohydrate, protein, zinc, and vitamin C contents in the food bars made of this flour need to be increased. Therefore, sword bean (Canavalia ensiformis) flour is chosen to produce better nutritional outcomes among under-five children.

The food bar is an appropriate type of emergency food for disaster refugees because it can be produced from various local food ingredients and its solid, compact form makes it easier to distribute to the refugees. It also contains high carbohydrates and proteins and can be directly consumed without processing them first. In addition, food bars are more resistant to pressure than biscuits or cookies because of their semi-wet nature [12]. A study on food bars made of *lindur* fruit (*Bruguiera gymnorrhiza*) flour, broccoli flour, and soybean flour was conducted in 2020. A total of 33 older people refugees affected by a flood in Depok City consumed the food bars for 15 days. The weight of these older people was found to increase by 0.2 kg during this period [10].

This study explored an EFP in the form of food bars made of a mixture of *api-api* mangrove flour and sword bean. *Api-api* mangrove was selected because it is rarely studied compared with *lindur* mangrove. This type of mangrove also contains carbohydrates, albeit in a slightly lower amount [13]. One hundred grams *of api-api* mangrove contains 21.43% carbohydrates, 10.4% protein, 0.04% fat, and 22.24/mL of vitamin C [14]. Meanwhile, in 100 g of sword bean beans, there are 55 g of carbohydrate, 24 g of protein, and 3 g of fat, producing 332 kcal of energy [15]. *Api-api* mangrove flour can be used as a substitute for wheat flour in making biscuits [11]. However, the use of *api-api* mangrove flour as a raw material for food products, such as biscuits and cookies, is still uncommon. Under-five children met their total calorie needs by consuming the food bars three times a day [16].

## Materials and methods

### Study design

A non-randomized pre-post intervention study was conducted in 34 under-five children affected by landslides disaster as the research subject [17]. The intervention group came from three neighborhoods (1,2,3) and the control group taken from other three neighborhoods (4,5,6).

### Ethical approval

Ethical clearance was obtained from the Ethics Commission for Health Research and Development (KEPPK) of the Sint Carolus, College of Health Sciences, Jakarta (No.089/KEPPKSTIKSC/VII/2022). All selected mothers of under-five children subjects signed a written informed consent form before fully participating in this study in the early August 2022. The signing was performed during the introductory activities of this study and witnessed by the integrated health post of under-five children (*posyandu)* cadres and research team.

### Population and subject

The study population consisted of under-five children who were victims of the landslide disaster in Cihanjuang Village, Cimanggung Sub-district, Sumedang District, West Java Province, Indonesia. The list of under-five victims of landslides disaster taken from the Cihanjuang Village Chief. The inclusion criteria for under-five children were aged between 12 and 59 months; male and female; living in Cihanjuang Village of Sumedang District; affected by the landslide disaster; had malnutrition, normal, or overnutrition status; not suffering from infectious diseases; and willing to avoid consumption of any snacks other than the food bars and plain water. The minimum sample size was calculated using the hypothesis test of paired mean difference formula [18]. This was based on the weight gain of 0.4 kg among under-five children with a standard deviation of 1.9 [19]. This study used a two-sided significance level of 0.05. Thus, a sample size of n = 15 was needed to reach 90% statistical power.

### Subject recruitment

The number of under-five children who fully participated until the end of the study was 35 subjects. Out of 35 under-five children subjects at the start of study, there were 17 subjects in the intervention and 17 subjects in the control group who fully participated until finished. One subject was excluded from the study because he getting bored with food bars and stop eating it (Figure 1).

**Fig 1.**
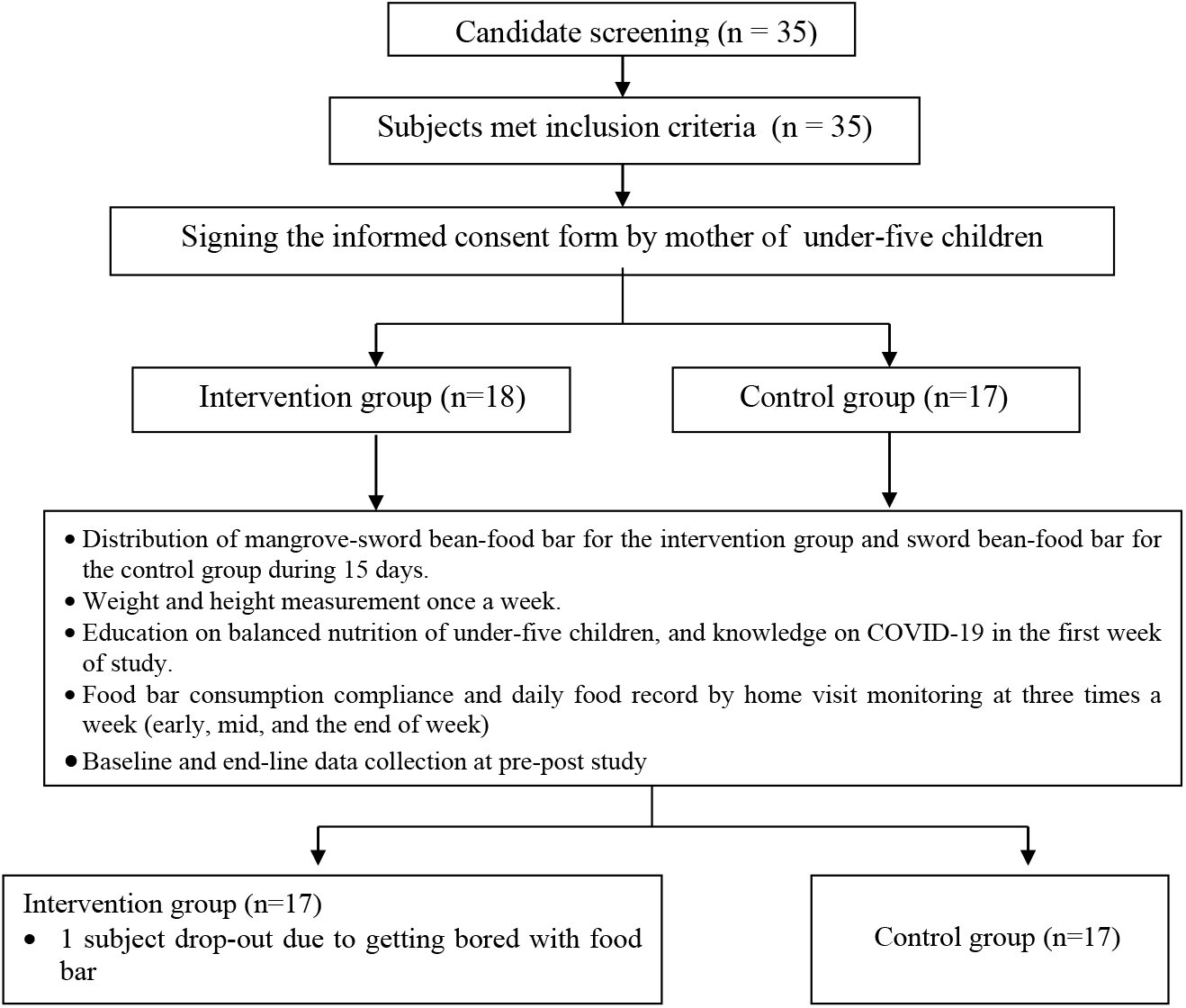
Research scheme of under-five children

The name list of under-five children landslide victims was obtained from the head of the integrated health post for under-five children (*posyandu*). The anthropometric measurements were conducted at the *posyandu* locations to collect the initial nutritional status data. The prospective subject were also invited to attend the dissemination to introduce the 15-day study [20].

### Nutrition intervention and follow-up

The under-five children in the intervention group received mangrove-sword bean-food bar during 15 days, which was calculated starting from the evacuation day as defined by the UNHCR [20]. Each day, the subject in the intervention group had 50 g food bars as snacks.

Every 50 g of food bar contains 234.2 kcal of energy, 24.7 carbohydrates, 5.8 g of protein, and 12.5 g of fat (Table 1).

**Table 1.** Nutrient content per 50 g of food bar

| Type of food bar | Energy (kcal) | Carbohydrate (g) | Protein (g) | Fat (g) |
| --- | --- | --- | --- | --- |
| Mangrove sword bean | 234.2 | 24.7 | 5.8 | 12.5 |
| Sword bean | 230.4 | 26.5 | 3.9 | 12.1 |

Source: Saraswanti Laboratory Bogor, 2022

The baseline questionnaire was used to collect data, and data were collected by enumerators by interviewing all mothers of under-five children to obtain an overview of the socio-demographic characteristics of mothers of under-five children (age, marital status, highest formal education, occupation, and status of cohabitation at home), as well as the characteristics of the under-five children (age and sex). The questionnaire also assessed the knowledge of balanced nutrition (healthy food menu; benefits of carbohydrates, proteins, vitamins, and minerals; definition of healthy food; definition of balanced nutrition; the need to measure weight; definition of stable or reduced weight with its causal factors and impacts; and efforts to increase weight).

Anthropometric data collection was performed three times: before, during, and after the study. The weight and height were measured using a digital scale and microtoise, respectively. Balanced nutrition and landslide disaster mitigation in the form of brochure and flipchart developed by the research team. The nutrition education was conducted in the first week for mothers of under-five children in the intervention group. The education for mothers of the under-five children was delivered in Indonesian. At the end of the study, endline data collection was performed to assess the extent of changes in balanced nutrition as well as the macronutrient intake, after education was provided.

### Hedonic/organoleptic test of food bar

Hedonic test was undertaken before mangrove-sword bean-food bar used as the EFP for the under-five children. The objective of test was to determine the preference of food bar: api-api mangrove-sword bean blends. Twenty mothers of under-five children who were selected from different area with the study sites designed as a untrained panelist in the organoleptic test. In the case of untrained panelists, 10–20 people are preferable [21]. All semi-trained panelists did not participate in the research subjects to avoid bias. They recognized the quality of food bar (color, aroma, taste, and texture) which can influence their compliance liking and acceptance the food bar. Aroma, texture, color, and taste of food bar were rated using a four-point hedonic scale ranging from 1 (very dislike), 2 (dislike), 3 (likes), and 4 (very like) [22].

### Cookie consumption compliance monitoring

Cookie consumption of subjects was monitored through three times home visits a week and anthropometric data measurement at *posyandu* once a week during the study. Data on food consumption, cookie distribution, the remaining number of cookies, and cookie consumption aftermath were recorded at each home visit. In the beginning, one subject felt cookies was hard. However, after the mother was explained by the enumerators about the side effects appeared as a form of early adaptation then her children continued it. Compliance level to taking the food bar in the under-five children was pretty good.

Food consumption record data were collected by trained enumerators during home visits, which were scheduled three times a week while distributing the food bars. The monitoring of food bar consumption compliance was carried out through the collection of anthropometric data on subjects at the first week and the second week of study, as well as by recording 24 hours/3 days/week food recalls.

### Food bar cooking process

The *api-api* mangrove sword bean food bar (Figure 2) is made from *api-api* mangrove flour with the addition of melted butter, chicken eggs, refined white sugar, a small amount of wheat, and chocolate/strawberry/orange pasta. The equipment used for making the food bar consisted of a digital oven, mixer, 20 cm × 15 cm aluminum brownie mold, digital scale, food bar dough cutting knife, and dough grinder. The steps for making food bars as follows:

1. Melt the butter and mix it with refined white sugar. A mixer was used to mix the dough for 5 minutes until it was homogeneous.
2. Add the *api-api* mangrove flour gradually into the mixture, followed by sword bean flour, egg yolks, and pasta.
3. Stir the dough gently with a large wooden spatula until it reached a homogenous consistency.
4. Pour the dough into a brownie mold and baked in a digital oven for 30 minutes at a temperature of 180°C.

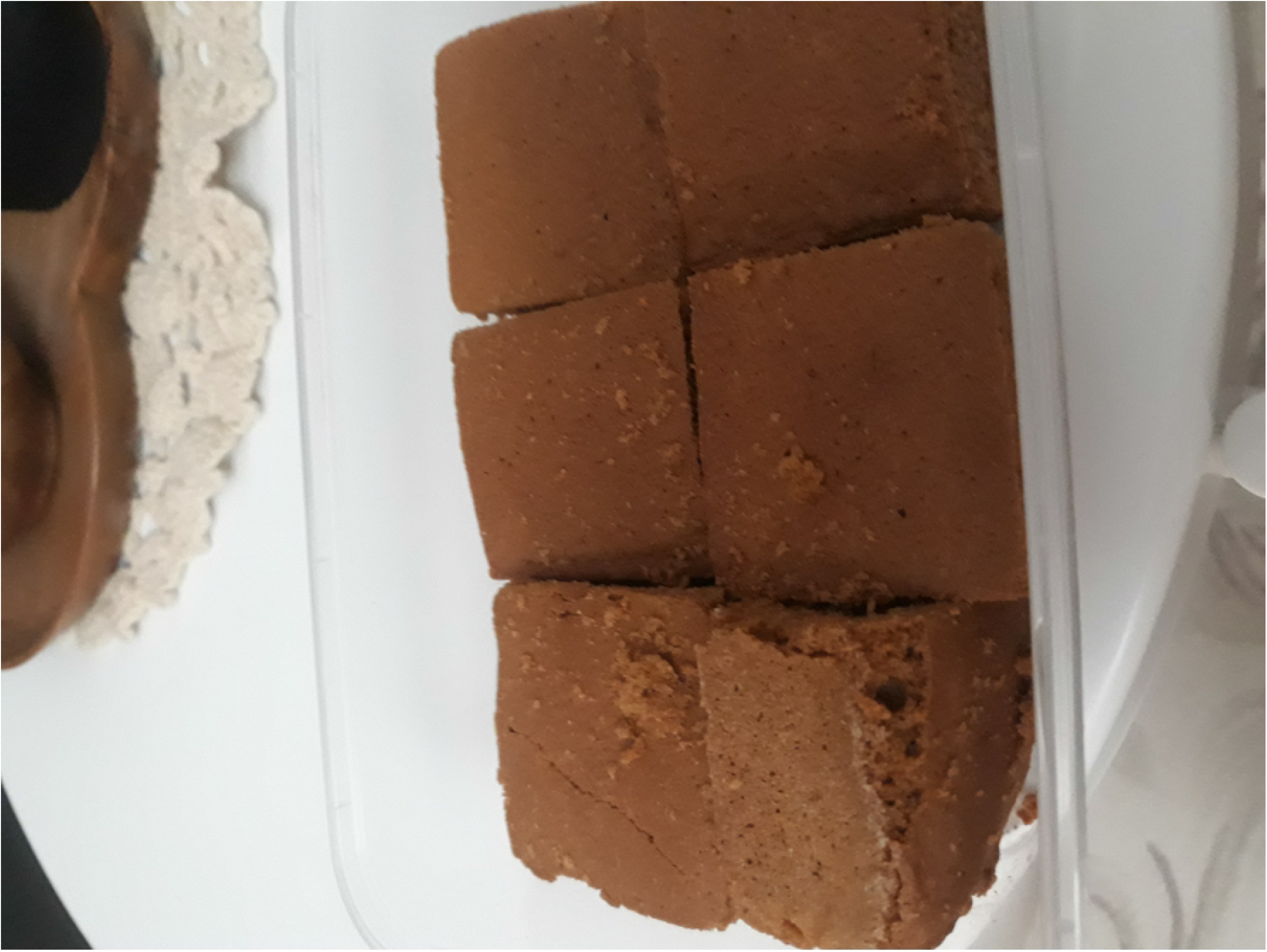

### Data analysis

Univariate analyses was performed to obtain the mean values of food bar organoleptic test; socio-demographic characteristics, the nutritional status, and knowledge of balanced nutrition and landslide disasters of mothers of the under-five children using SPSS version 22 software. The 2-weeks food consumption data analyzed by The Nutri Survey Program [23]. The food data consumption data analyses aimed to measure change of macronutrient intake (energy, carbohydrate, fat, and protein) at pre-post study. Bivariate analysis with a statistical paired t-test was used to assess changes in mean weight, Weight for Age Z-score (WAZ), macronutrient intake (energy, carbohydrate, protein, and fat), and balanced nutrition knowledge for under-five children.

## Results

The study assessed the mangrove-sword bean-food bar preference using four parameter: color, aroma, taste, and texture. Mothers of under-five children more liked the mangrove-sword bean food bar from color, aroma, taste, and texture than sword bean food bar (Table 2).

**Table 2.** Organoleptic test of mangrove-sword bean and sword-bean food bars by mothers of under-five children

| Parameter | The mean value of food bar |  |
| --- | --- | --- |
|  | Mangrove -sword bean food bar | Sword bean food bar |
| Color | 3.10 ± 0.60 (like) | 3.10 ± 0.60 (like) |
| Aroma | 3.05 ± 0.45 (like) | 2.15 ± 0.14 (dislike) |
| Taste | 3.04 ± 0.32 (like) | 2.20 ± 0.45 (dislike) |
| Texture | 3.14 ± 0.23 (like) | 2.10 ± 0.12 (dislike) |

1=very dislike; 2=dislike; 3= likes; 4 = very like

The socio-demographic characteristics of under-five children subject in this study were divided into maternal (marital status, age, final education, employment status, and cohabitation status at home) and under-five children characteristics (sex and age) in the intervention and control groups (Table 3). Almost all the mothers of the under-five children in both groups were married with age ranging from 20 to 29 years old. Most mothers of the under-five children graduated from junior high school and were unemployed. Most of the subject lived in nuclear family with the arrangement of father, mother, and son/daughter was mostly identified as living in the same house with the subjects. More girls participated in this study than boys. The mean age of under-five children in both groups was 36 months, with the mean age of intervention group slightly lower than that of the control group.

**Table 3.** Socio-demographic characteristics of under-five children subjects

| Variable |  | Control |  | Intervention |  | Total |  |
| --- | --- | --- | --- | --- | --- | --- | --- |
|  |  | n | % | n | % | n | % |
| *Characteristic of family* |  |  |  |  |  |  |  |
| Marital status |  |  |  |  |  |  |  |
|  | Married | 16 | 94.1 | 17 | 100.0 | 33 | 97.1 |
|  | Widow/widower | 1 | 5.9 | 0 | 0.0 | 1 | 2.9 |
| Age of mother (years old) |  |  |  |  |  |  |  |
|  | Mean ± SD | 30.7 ± 6.8 |  | 30.6 ± 7.5 |  | 30.6 ± 7.0 |  |
|  | 20–29 | 8 | 47.1 | 9 | 52.9 | 17 | 50.0 |
|  | 30–39 | 7 | 41.2 | 7 | 41.2 | 14 | 41.2 |
|  | 40–50 | 2 | 11.8 | 1 | 5.9 | 3 | 8.8 |
| Mothers' education |  |  |  |  |  |  |  |
|  | Graduated from junior high school or less | 11 | 64.7 | 8 | 47.3 | 19 | 55.9 |
|  | Graduated from senior high school | 5 | 29.4 | 8 | 47.1 | 13 | 38.2 |
|  | Academy / bachelor | 1 | 5.9 | 1 | 5.9 | 2 | 5.9 |
| Working status of mother |  |  |  |  |  |  |  |
|  | No | 11 | 64.7 | 11 | 64.7 | 22 | 64.7 |
|  | Yes | 6 | 35.3 | 6 | 35.3 | 12 | 35.3 |
| Living arrangement |  |  |  |  |  |  |  |
|  | Family (father, mother, children) | 15 | 88.2 | 17 | 100.0 | 32 | 94.1 |
|  | Join with another family | 2 | 11.8 | 0 | 0.0 | 2 | 5.9 |
| *Characteristic of under-five children* |  |  |  |  |  |  |  |
| Sex |  |  |  |  |  |  |  |
|  | Female | 12 | 70.6 | 8 | 47.1 | 20 | 58.8 |
|  | Male | 5 | 29.4 | 9 | 52.9 | 14 | 41.2 |
| Age (month) |  |  |  |  |  |  |  |
|  | Mean ± SD | 38.4 ± 12.1 |  | 33.9 ± 14.3 |  | 36.1 ± 13.2 |  |

Table 4 displays the anthropometric data (weight and WAZ score) of under-five children at before and after the intervention. At the end of the study, the under-five children in the intervention group had a slightly higher weight gain than the control group. The under-five children’s weight in each group demonstrated a significant difference between before and at the end of the study (p <0.001). However, this was not the case when the weight was compared between groups (p > 0.05). The intervention group are likely to experience a WAZ score greater as a result of a food bar supplementation intervention compared with the control group. The difference in mean WAZ score change between the intervention and control groups was statistically significant (p=0.023).

**Table 4.** The mean difference of weight and WAZ score of under-five children at pre-post study

| Indicator | Control |  | Intervention |  | Different | *p* |
| --- | --- | --- | --- | --- | --- | --- |
|  | Mean | SD | Mean | SD |  |  |
| Weight (kg) |  |  |  |  |  |  |
| At pre/before | 13.18 | 2.83 | 12.01 | 2.17 | 1.17 | 0.186 |
| At post/after | 13.42 | 2.82 | 12.34 | 2.14 | 1.09 | 0.215 |
| Different | 0.24 | 0.17 | 0.32 | 0.14 | 0.08 | 0.138 |
| 95% CI of different | (0.13 – 0.33) |  | (0.25 – 0.39) |  |  |  |
| *p* * | **<0.001 |  | **<0.001 |  |  |  |
| WAZ score (Weight for Age) |  |  |  |  |  |  |
| at pre/before | -0.68 | 1.27 | - 0.32 | 1.04 | -0.36 | 0.373 |
| at post/after | -0.88 | 1.19 | -0.01 | 0.91 | -0.87 | **0.023 |
| Different | -0.19 | 0.82 | 0.31 | 0.73 | 0.51 | 0.067 |
| 95% CI of different | (-0.18 – 0.57) |  | (-0.11 – 0.74) |  |  |  |
| *p* * | 0.288 |  | 0.137 |  |  |  |

#*p*: independent *t*- test

Table 5 displays the mean difference of macronutrients intake, score of knowledge on the under-five children balanced nutrition, and food bar total consumption. At the end of study, those in the intervention group showed significantly greater increment in fat intake and food bar total consumption. There were significant result was found for under-five children mothers’ knowledge on the under-five children balanced nutrition in the two groups at post study.

**Table 5.** The mean of macronutrient intake of under-five children and balanced nutrition knowledge of mothers

| Indicator | Control | Intervention | *p* |
| --- | --- | --- | --- |
| | Mean ± SD | Mean ± SD | |
| Energy (kcal) |  |  |  |
| At 1<sup>st</sup> week | 1106.9 ± 253.2 | 977.3 ± 223.5 | 0.124 |
| At 2<sup>nd</sup> week | 1156.5 ± 310.7 | 1106.9 ± 253.2 | 0.193 |
| *p* * | 0.293 | 0.338 |  |
| Carbohydrate (g) |  |  |  |
| At 1<sup>st</sup> week | 149.9 ± 43.9 | 130.9 ± 34.4 | 0.171 |
| At 2<sup>nd</sup> week | 162.5 ± 57.5 | 134.5 ± 32.7 | 0.093 |
| *p* * | 0.252 | 0.643 |  |
| Protein (g) |  |  |  |
| At 1<sup>st</sup> week | 34.3 ± 9.9 | 30.1 ± 11.1 | 0.253 |
| At 2<sup>nd</sup> week | 37.9 ± 15.1 | 32.0 ± 8.9 | 0.180 |
| *p* * | 0.187 | 0.496 |  |
| Fat (g) |  |  |  |
| At 1<sup>st</sup> week | 39.9 ± 11.7 | 33.7 ± 10.6 | 0.107 |
| At 2<sup>nd</sup> week | 45.7 ± 21.8 | 55.6 ± 9.4 | **0.04 |
| *p* * | 0.601 | 0.111 |  |
| Total consumption of food bar (g) | 588.2 ± 158.6 | 650.5 ± 100.9 | **0.045 |
| Balanced nutrition knowledge of mothers |  |  |  |
| Pre-study | 42.2 ± 6.9 | 43.5 ± 16.1 | **< 0.001 |
| Post-study | 62.2 ± 13.3 | 65.7 ± 19.4 | **< 0.001 |

#*p*: independent *t*-test

## Discussion

Nutrition management activities during and after the reconstruction and rehabilitation stages of a disaster aim to improve and maintain the nutritional status of victims of natural disasters [24]. The nutritional requirements of under-five children during and after the disaster have not been specifically handled, and there is no specific food available for under-five children. In addition, the amount and type of food available for disaster victims are not yet appropriate and cannot meet the nutritional needs of under-five children [25]. Under-five children are the most vulnerable group affected by disasters, and the provision of emergency food is not only needed to meet nutritional feasibility, but also to meet nutritional needs. Providing nutritious food and nutrition education can lead to positive results in preventing malnutrition among under-five children after natural disasters while also increasing food security [26].

Nutrition management after a disaster through surveillance activities in the form of anthropometric assessment of under-five children and nutrition awareness activities, including nutrition education and consumption of food bars as a follow-up or response to the information obtained from public health service activities, can improve and maintain the nutritional and health status of disaster victims [27]. After a natural disaster strikes, there is an increase in the number of under-five children who are undernourished and stunted, emphasizing the need for nutrition and education interventions. Integrated nutritional interventions performed in collaboration with local health services, such as *posyandu* for under-five children can result in optimum results. The readiness of individuals and institutions to build local safety culture can increase post-disaster resilience. Food security needs to be included in the recovery efforts to maintain the nutritional and health status of group that are vulnerable to disasters, such as under-five children. The Sendai Framework has prioritized under-five children group in reducing disaster risk [28]. Increased active community participation through compliance with emergency food consumption during post-disaster rehabilitation plays a role in the post-disaster recovery process.

The increase in weight of under-five children measured in this study is 0.2 kg, which supports the findings of several studies reviewed by Pradhan et al. in 2016. Nutritional supplementation of under-five children after a disaster can improve the nutritional status of children who experience undernutrition, have short stature, and are underweight [29]. Breastfeeding for up to 2 years for under-five children after the Palu earthquake has been shown to reduce the incidence of stunting [28]. The current study described an increase in weight and WAZ score in 15 days, but are unable to prove any significant differences in the nutritional status of under-five children based on the maternal socio-demographic characteristics, knowledge of balanced nutrition, and the macronutrient intake of the under-five children, except fat intake. This may be due to the relatively short intervention of 15 days; small sample size (18 under-five children); mean of under-five children with normal nutritional status (WAZ), and the consumption of food bars as snacks.

The total consumption food bar and fat intake for under-five children influenced changes in the weight and Z-score of W/A in the present study based on the comparison of the pre-post study values. The daily consumption of macronutrients for under-five children after a disaster met 50% of the RDA (Recommended Daily Allowance) with the daily consumption of 50 g of mangrove food bars containing 234.2 g of energy, 24.7 g of carbohydrate, 12.5 g of fat, and 5.8 g of protein. The finding of current study contradicts the findings demonstrated in a previous study that showed a decrease in the prevalence of under-five children who are underweight and have short stature after a disaster [30]. The nutritional management for under-five children after disaster should integrated nutrition interventions with public health promotion activities such as nutrition counselling campaign or education [31].

Knowledge of balanced nutrition of mothers of under-five children differed significantly at the end of the study. These findings are in line with three studies that proved nutrition education improves the nutritional status of thin, short, and under-five children with anemia [32,33,34]. On the other hand, low maternal knowledge of balanced nutrition affects the pattern of feeding practices and efforts to meet the nutrition requirements of under-five children [35]. Nutrition education is effective in improving nutritional knowledge and eventually increasing nutritional status of under-five children.

The present study has some limitations. First, the small sample size for this study was recruited from one selected sub-village only so it cannot be generalized for Sumedang District. Second, the short duration in the two weeks might influence the change weight and WAZ to be not optimal. For future research, it is recommended a similar study measuring height and WHZ of under-five children with big sample size (for example 40 subjects) and long duration (for example 3 months) combining with the nutrition counselling using the control group as the comparison.

## Conclusion

Although, there is no significant increase in the WAZ score, but there is a significant difference in the weight of under-five children after consuming the *api-api* mangrove and sword bean food bar. Thus, this food bar is still suitable for emergency food alternatives during and after a disaster due to its nutrient contents and form. Nutrition education is also an important aspect of nutrition provision to disaster refugees, especially for vulnerable groups, such as under-five children. Further research with a bigger sample size and a longer period is needed to produce evidence on the impact of emergency food, such as food bars on the nutrition status of disaster refugees, especially for vulnerable groups such as older people and pregnant women. These further studies also need to involve different types of nutritional status, including underweight, stunted under-five children or underweight older people.

## Data Availability

All relevant data are within the manuscript and its Supporting Information files.

## Acknowledgements

The authors thank all parties who have supported the smooth implementation of the study, including all the older people and mothers of the under-five children participating in this study, as well as cadres of the Integrated Health Post (*posyandu*) for older people and under-five children in Cihanjuang Village, Cimanggung subdistrict, Sumedang District and all field enumerators.

